# What have we learned so far about COVID-19 volunteering in the UK? A rapid review of the literature

**DOI:** 10.1101/2020.11.22.20236059

**Authors:** Guanlan Mao, Maria Fernandes-Jesus, Evangelos Ntontis, John Drury

## Abstract

**Background:** Community engagement and volunteering are essential for the public response to COVID-19. Since March 2020 a large number of people in the UK have been regularly doing unpaid activities to benefit others besides their close relatives. Although most mutual aid groups emerged from local neighbourhoods and communities, official public institutions also fostered community volunteering, namely through the community champions scheme. By considering a broad definition of COVID-19 volunteering, this article describes a systematic review of the literature focused on two broad questions: What have we learned so far from COVID-19 volunteering both at the UK national level and the more local community level? What have we learned from engagement with local communities and community champions during the COVID-19 period?

**Methods:** A rapid review of the literature in peer-reviewed databases and grey literature was applied in our search, following the PRISMA principles. The search was conducted from 10 to 16 of October 2020, and sources were included on the basis of having been published between January and October 2020, focusing on COVID-19 and addressing community groups, volunteering groups, volunteers, or community champions in the UK.

**Results:** After initial screening, a total of 40 relevant sources were identified. From these, 28 were considered eligible. Findings suggest that food shopping and emotional support were the most common activities, but there were diverse models of organisation and coordination in COVID-19 volunteering. Additionally, community support groups seem to be adjusting their activities and scope of action to current needs and challenges. Volunteers were mostly women, middle-class, highly educated, and working-age people. Social networks and connections, local knowledge, and social trust were key dimensions associated with community organising and volunteering. Furthermore, despite the efforts of a few official public institutions and councils, there has been limited community engagement and collaboration with volunteering groups and other community-based organisations.

**Conclusions:** We identified important factors for fostering community engagement and COVID volunteering as well as gaps in the current literature. We suggest that future research should be directed towards deepening knowledge on sustaining community engagement, collaboration and community participation over time, during and beyond this pandemic.

## Background

In the absence of safe and effective vaccines or drugs, communities all over the world depend heavily on behavioural measures, such as physical distancing, curtailing of social gatherings, face coverings, and regular hand-washing to contain the spread of COVID-19 and prevent their healthcare systems being overwhelmed (1). Such measures have been frequently used to reduce the risk of infectious diseases spreading in previous pandemics (2, 3), and there is evidence already that they are helping contain the COVID-19 pandemic (4, 5). Among the most difficult of these non-pharmaceutical interventions is self-isolation, with UK surveys estimating that less than 20% of those required reported doing so for the full 10-14 days^1^ (3, 6, 7, 8, 9). To ensure that people follow COVID-19 measures, police enforcement and fines for failure of self-isolation have been employed (11, 12). However, data collected in the UK shows that these policies may lead to a decrease in self-reporting (13). Instead of coercion, scientific advisors have called for ‘supported isolation’, which includes providing appropriate accommodation, domestic assistance, and financial support (6, 14). The latter is more in line with World Health Organization (WHO) recommendations to avoid coercion in public health (15, 16) and to adopt instead a community engagement approach (17, 18). Community engagement in public health refers to the involvement and participation of individuals, groups and structures in the decision-making, planning, design, governance and delivery of services (18, 19). Past pandemics have provided sufficient evidence that community engagement is crucial in fostering public health in pandemic conditions, with community action groups playing a key role in reducing contagion (17, 20, 21).

The COVID-19 pandemic has provoked a remarkable surge in volunteering and community support around the world (22, 23). Prominent manifestations of this outpouring of community spirit within the UK include the rise of so-called mutual aid groups, volunteer-led initiatives where individuals from a particular area group together to meet community needs without the help of official bodies (24). Over 4000 such groups have formed over the course of the pandemic, with as many as three million participants (25). On a national level, the NHS volunteer responders scheme was able to recruit over 750,000 people in four days, three times the initial target (26). Additionally, some local authorities further promoted community champions programmes during the pandemic (27). Community champions are trained and supported volunteers who help improve the health and wellbeing of their communities. They share information, motivate and empower people to get involved in health-promoting activities, create groups to meet local needs, and direct people to relevant support and services (28).

The benefits of such forms of community volunteering seem to go beyond the context of the pandemic conditions, with previous research suggesting that volunteering may benefit mental health and survival, fostering well-being and life satisfaction (29), as well as sense of belonging (30). In this study, we focus on a broad definition of COVID-19 volunteering, to capture the multiple ways people engaged in community support and mutual aid groups, as well as in community champions programmes. Volunteering is here considered as any unpaid activity that involves spending time carrying out tasks that aim to benefit people other than close relatives (31). With this in mind, this article reports a rapid review of the literature that addresses two broad questions: 1) What have we learned so far from COVID-19 volunteering both at the UK national level and the more local community level? 2) What have we learned from engagement with local communities and community champions during the COVID-19 period? Answering these questions may illuminate political, organisational and psychological aspects of COVID-19 volunteering.

## Methods

A rapid review of literature was applied in our search. Rapid reviews are a useful form of producing information in a timely manner (32). A multi-faceted approach, following the PRISMA principles (33), was adopted in order to scope the rapidly evolving literature base for the topic. Searches were conducted in peer-reviewed databases and grey literature. The inclusion of grey literature in public health reviews can help advance the understanding of what and how interventions are being implemented (34). As grey literature is not controlled by commercial publishing organisations, the information is published when a particular phenomenon is occurring. This is particularly useful for applied researchers and practitioners, in disasters and pandemic conditions such as COVID-19, where relevant and timely information is urgently needed.

### Search strategy

The search was applied to six databases: ScienceDirect, University of Sussex Library, APA Psycnet, Wiley Online Library, PubMed and SocArXiv. These databases were selected based on their coverage of the topic. For grey literature, a search was applied for relevant think tank, governmental, and third sector organisation websites. Academic websites, blogs and research networks were also considered in our search. Finally, we conducted a web search via Google Advanced.

The search was conducted between October 10 to 16, 2020, using the following keywords: “COVID-19” OR “Coronavirus”, AND “volunteering” OR “mutual aid” OR “community”, “community engagement”, OR “community champions”. Records were included if they were published between January and October 2020, focused on COVID-19 and addressed community groups, volunteering groups, volunteers, or community champions in the UK. We considered many source types, including published peer reviewed articles, reports, briefings, blogposts, newspaper articles, and online media relevant to research questions. Only English sources were considered.

## Results

After initial screening, a total of 40 relevant sources were identified, including two from published literature databases, 29 from governmental and third sector organisation websites, think-tanks, six from Google Advanced, and three from academic websites, blogs and research networks. After all of these sources were assessed for eligibility, 28 were included in the final qualitative synthesis (Fig. 1).

**Fig 1.**
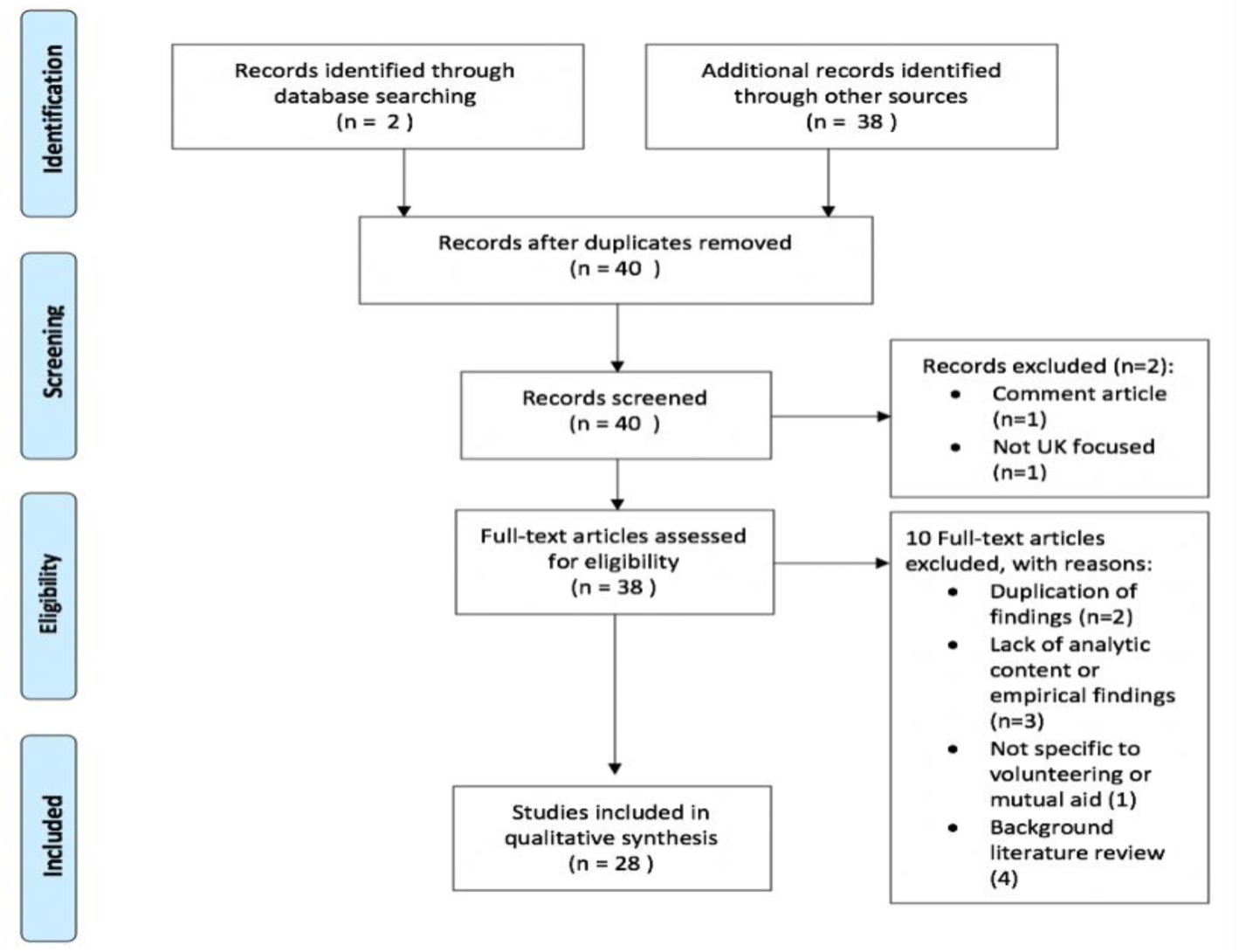
Flow Diagram representing the selection process of articles

Table 1 reports the characteristics of the sources analysed, including the type of source, the setting, the sample size (if applicable), the study design, the process followed in data collection, and a short summary of the major findings. Most eligible sources identified can be considered grey literature and were produced by civil society organisations. From the 28 sources included in the qualitative analysis there were: 13 reports, three briefings, five blogposts, two newspaper articles, two websites entries, one handbook, and two peer reviewed journal articles. 15 sources used primary data, 11 secondary and two sources were based on both types of data. A few sources were based on large surveys with volunteers, but the majority focused on qualitative forms of enquiry (e.g., interviews, conversations) with volunteers, stakeholders, or organisations. Most of the sources focused on the national context or on a large set of regions. To address our two research questions, in the following sections we will describe the qualitative findings of this review by focusing on the six major topics: volunteering activities; models of volunteering; volunteer profiles; successes, challenges, and determinants of effectiveness; collaboration with local communities; and consulting local communities.

**Table 1.**
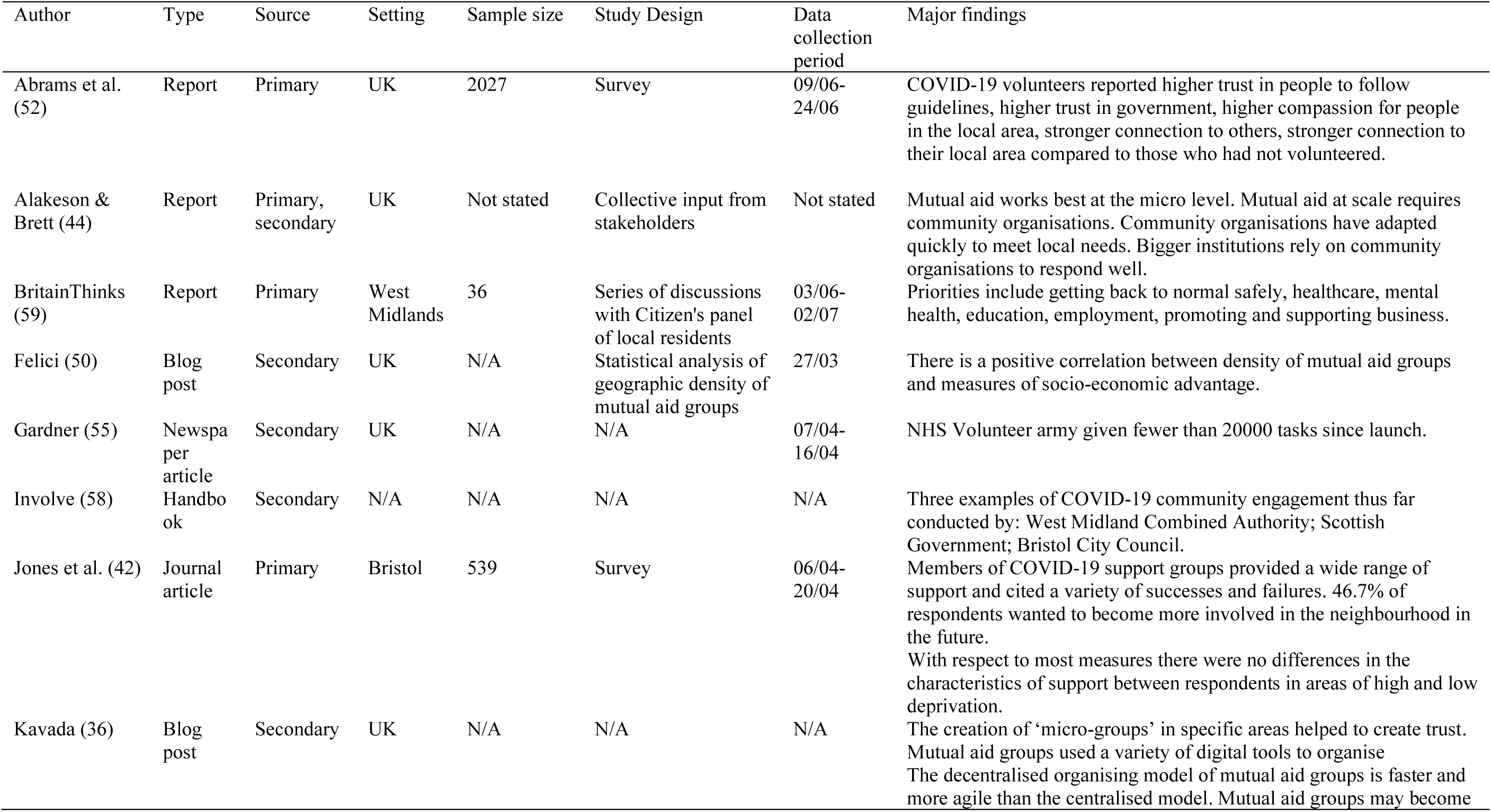
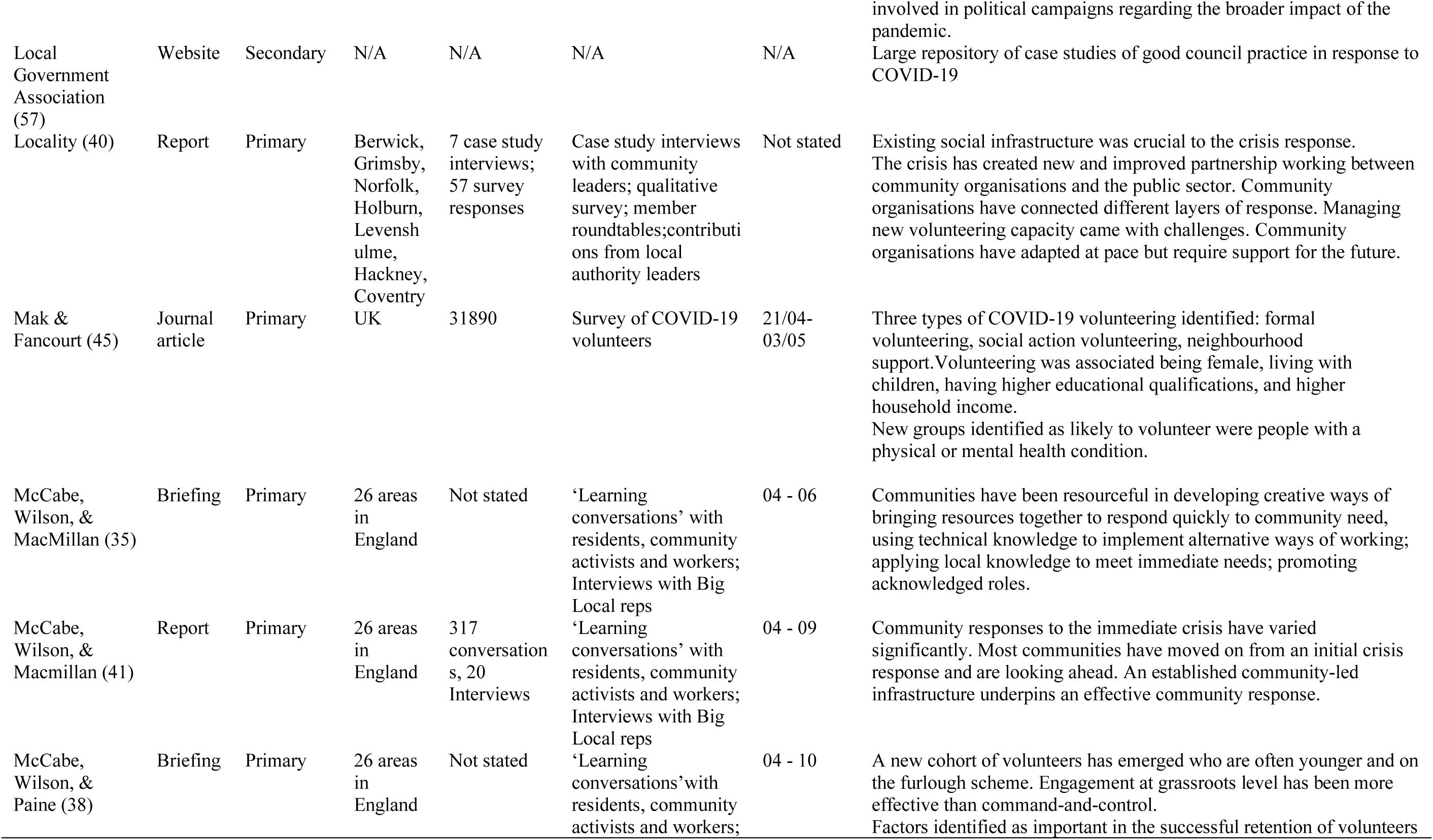
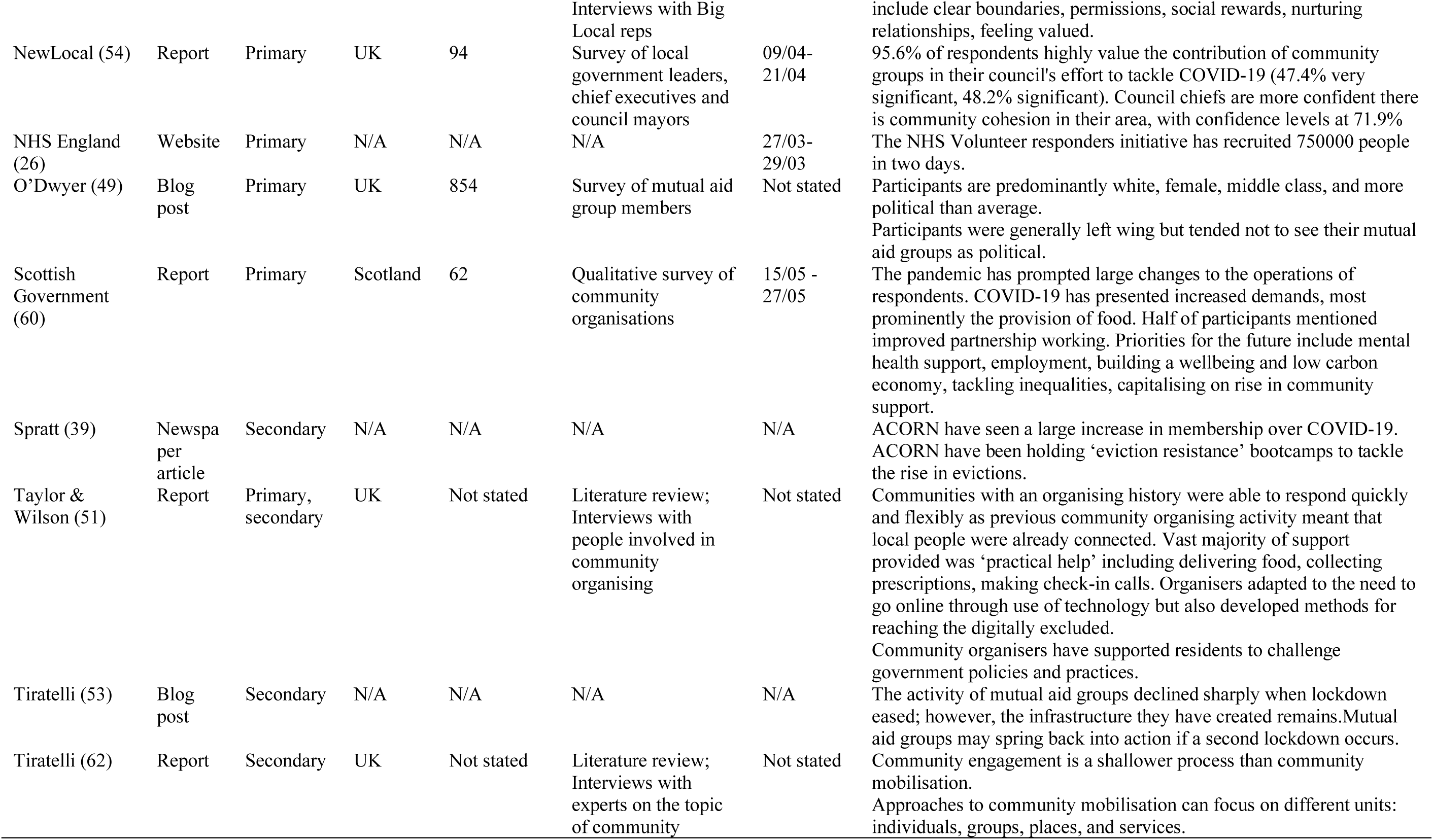
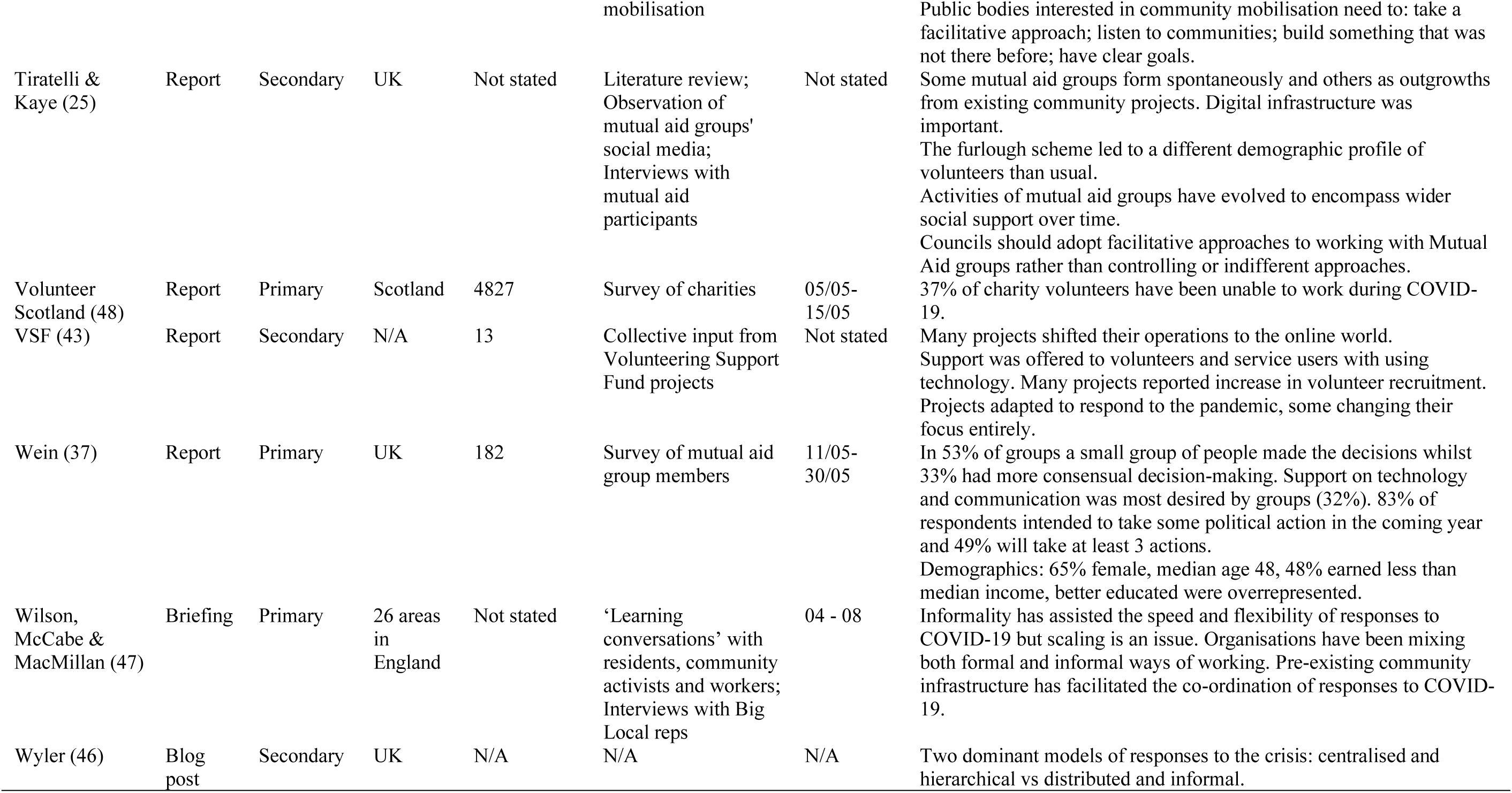
Characteristics of included sources

### Volunteering activities

Studies reviewed indicate that delivery of essentials such as food and prescriptions dominated early efforts in COVID-19 volunteering (35). A second type of activity which became increasingly common as the first ‘lockdown’ wore on was combating social isolation through activities such as provision of arts and crafts packs, telephone support, and online social activities (35). After the first ‘lockdown’ (23^rd^ March – June-July 2020), there was an increasing shift towards volunteering activities that address the wider impact of the pandemic on other areas such as employment, social benefits, mental health, domestic abuse, and homelessness (35, 36). There is also some evidence that COVID-19 volunteers became involved in wider political campaigns (36, 37). Wein (37) found that 83% of mutual aid participants intended to take part in some form of political action in the coming year, with 64% likely to sign petitions and 47% expecting to contact a politician. Information about the Black Lives Matter movement has reportedly been circulating in WhatsApp groups, whilst activists associated with mutual aid groups staged an action outside the house of Dominic Cummings (36). In one case study described by McCabe et al. (38), volunteers who witnessed the living conditions of those they were helping developed a collective action approach to addressing poor housing conditions. On an organisational level, ACORN, a community union which organised mutual aid networks around the country, has worked to divert many of its volunteers from community support to eviction resistance campaigns (39).

In many cases, existing voluntary organisations and projects adapted their services by transferring to digital infrastructure, often at a rapid pace (40). A plethora of digital tools were put to use. Whilst WhatsApp was one of the most popular organising platforms, some groups adopted more streamlined services such as Slack. Platforms such as Zoom and Skype were used for group calls; Google Docs for meeting minutes; and Google Sheets for compiling databases of volunteers and requests (36). Other groups recreated their activities online, such as a weekly Facebook-based interactive youth club (41). Many groups sought to tackle the possibility of digital exclusion through physical methods such as mass leafleting (42). Other projects (for example Skills Enterprise in East Ham) offered digital training sessions, as well as providing tablets and phones to those on their programmes (40, 43). The crisis has demonstrated the adaptability and resourcefulness of volunteers and community organisations, who have effectively adjusted to changing conditions as the UK continues to pass through different phases of the pandemic. Importantly, their activities expose the insufficiency of public services (25), as these volunteers are serving to meet needs which are otherwise unmet by public services.

### Models of volunteering

The onset of lockdown saw an outpouring of community spirit and voluntarism, channelled in a huge variety of ways (35). Whilst in some areas volunteering activity surfaced spontaneously, in other areas this activity emerged as an outgrowth of existing networks, community projects, and organisations (25). In many cases such organisations shifted their activities rapidly to COVID-19, mobilising volunteers and relationships with other local groups to create local support schemes (40). For example, Homebaked in Anfield, a community bakery, closed down much of its traditional operations and started baking 50 to 70 loaves a day, which it provided to the local food bank and community centre (44). Focusing precisely on the models and predictors of volunteering, a large survey with 31,890 adults in the UK identified three types of volunteering during COVID-19 (45). The first, ‘formal volunteering’, included volunteering in formal and pre-existing structures and organisations. The second type, ‘social action volunteering’ was described as more oriented to broad fundraising and donation campaigns. Finally, ‘neighbourhood support’ involved providing support locally (e.g., shopping or cooking meals for others) (45).

Other authors have noted the emergence of two models of volunteering coordination during the pandemic (36, 46). On the one hand, a decentralised model, where information and decision-making are dispersed among members. On the other hand, a centralised method of command-and-control. These authors have generally argued for the superiority of the former model in terms of its speed, democratic nature, and ability to meet the needs of those excluded from other services (36, 46). Kavada (36) compared the model of mutual aid groups to the NHS volunteer responders service. The formal nature of the NHS scheme meant that the identities of all volunteers had to be carefully checked, leading to delays in assignment. Furthermore, the service only served UK inhabitants who registered as vulnerable, excluding those unwilling or unable to register formally (36). In contrast, mutual aid groups did not engage in verification of volunteers, and supported anyone who was self-isolating, allowing them to meet the needs of their communities more effectively (36). However, one common challenge reported by many mutual aid groups was a lack of leadership, where people were keen to offer services but were not willing to take the initiative (25). Equally, those engaged in more informal ways forms of supporting their neighbours also frequently reported the same challenge regarding reaching vulnerable groups, with help either lacking focus or being limited to those cases already known (35, 42). Furthermore, categorising volunteer activity as hierarchical or non-hierarchical, centralised or decentralised, formal or informal seems an oversimplification. In reality, most organisations combined elements of both approaches (47). Many groups preserve a ‘private layer’ of interaction for ‘core’ members and organisers (25), and group administrators were able to participate in a closed Facebook group to exchange tactics (36). Despite this caveat, sources reviewed suggest that the pandemic has prompted a qualitative shift in volunteering around the country, with traditional formal organisations such as charities losing a large bulk of their volunteers whilst informal associational models are thriving (48).

### Volunteer profiles

The conditions of the pandemic should arguably pose a challenge for volunteering efforts given its high risk to the elderly, normally the demographic most likely to volunteer regularly (31). However, the present circumstances appear to have led to the emergence of a new volunteer workforce. Results of large surveys (37, 49) suggest that the average age of COVID-19 mutual aid group members was 48. Another study (45) reported a slightly higher mean age (52 years old), concluding that older people were more likely to participate in volunteering than younger people, particularly in activities that involve providing local neighbourhood support. More generally, mutual aid groups appear to be concentrated in areas with large numbers of working-age people, a clear consequence of the government’s furlough scheme (25).Volunteers were also composed of more women than men (37, 49), especially in neighbourhood volunteering and social action volunteering (45). Whilst this is in line with general trends (31), it may also represent an extra caring responsibility at a time when women were already shouldering the burden of increased domestic labour.

There were also early indications that wealth and class played a role in participation. An analysis by Felici (50) of voluntary support networks across the UK revealed a positive correlation between the density of voluntary groups in an area, which is one of the manifestations of social capital, and measures of socioeconomic advantage, as well as well-being. In turn, Wein (37) argued that participants themselves are not necessarily wealthy. His survey indicates that 48% of volunteer households had an income of less than £30,000 and 30% above, compared to the national median of £29,600. Similarly, Mak and Fancourt (45) found that income predicted engagement in social action volunteering but did not predict other types of volunteering. However, it is important to remember that the resources and tactics available to these participants, and therefore the overall effectiveness of their participation, may not be the same. Indeed, a report by Taylor and Wilson (51) based on the experiences of community organisers found that whilst most affluent communities organise themselves, communities within more deprived areas often need more support but lack access to resources. In this regard, one participant in a mutual aid group from a poor rural area suggested that tactics such as crowdfunding would not be effective in rural areas (25). Nevertheless, Mak and Fancourt (45) found that whereas people who lived in rural areas were more likely to engage in formal and neighbourhood volunteering, there were no differences in terms of social action volunteering.

In terms of psychosocial and personality predictors, the only study that addressed these predictors found that personality traits (e.g., agreeableness, extraversion), but also social support and social networks were associated with engagement in all types of voluntary work during the pandemic (45). Interestingly, Abrams et al. (52) found that compared to people who had not volunteered, those who were volunteers during COVID-19 reported higher trust in people to follow guidelines, trust in government, and compassion for people in the local area. They also scored higher on connection to family, friends, colleagues and neighbours, and connection to their local area (52).

### Successes, challenges, and determinants of effectiveness

By delivering vital services to vulnerable individuals in the early days of lockdown whilst traditional public services struggled to respond effectively, mutual aid groups undoubtedly played a life-saving role in the UK’s COVID-19 response (25). Such groups have also generated new partnerships, networks and knowledge, which may serve as a long-term resource in the second wave (53). In terms of community and voluntary organisations generally, 95% of council leaders and chief executives saw community groups as being significant or very significant in their COVID-19 response (54).

However, volunteer groups have also faced many challenges. Many have found it hard to sustain the morale and enthusiasm of volunteers over time, with the activity of many groups declining sharply once lockdown started to ease (53). Other volunteering schemes found it hard to generate sufficient demand or faced high bureaucratic procedures that delayed their interventions (35). For example, the length of time it took for volunteers to hear back from the NHS Volunteer Responders Scheme caused initial enthusiasm to dissipate (40). Later data revealed that in the first week of the scheme, the 750,000 volunteers were given fewer than 20,000 tasks between them (55). By contrast, smaller mutual aid groups who attempted to scale up their operations beyond street level often found that they were lacking in organisation, coordination, local relationships, and trust (44).This was the case with a group formed in Dalston, London, which quickly attracted hundreds of volunteers but was unable to attract requests for support due to distrust from the local community (44).

In terms of sustaining volunteering, factors identified by groups as being important to successful retention of volunteers included: not asking volunteers to engage in activities they are uncomfortable with; allowing volunteers to say no; providing social rewards; nurturing relationships with volunteers; and recognising the contributions of volunteers (8). Moreover, a common theme emerging from the research reviewed is that effective and rich responses are underpinned by ‘community-led infrastructure’, understood as community leadership, trust, relationships with agencies, and access to funds (35). In particular, many community organisations have been able to play a coordinating role by providing smaller mutual aid groups with the infrastructure, systems, and resources required, as well as acting as a communication bridge between groups and local authorities (40). Local knowledge has also been important in responding to the needs of groups not covered by government schemes, such as homeless people or families with young children (47). For example, the Hastings Emergency Action Response Team (HEART) has been able to coordinate over 900 volunteers, using their local knowledge to identify needs (44). Additionally, the nationwide union ACORN was able to set up support systems in nine cities by mid-March. After years of organising and campaigning, ACORN already had an engaged existing membership in each city and well-developed organisational structures (51).

### Collaboration with local communities

The above discussion naturally raises the question of how authorities can best support local community-led infrastructures. In some cases, this was achieved through the COVID-19 Community Champion scheme (56, 57). These volunteers were given the latest information about Covid-19 and were asked to share this information in their community, whilst feeding back which communications are effective, and which are not (56). Councils which have successfully implemented the scheme include Newham Council, which have recruited more than 400 people to date (57). Champions receive messages through WhatsApp or email most days, including infographics which are available in a variety of languages. Among other things, they are given a badge and are included in a WhatsApp group so that they can share advice and support one another. Newham Council have now supported more than 30 other councils to develop their own programmes (57). Despite such successful case studies, as of the time of writing no systematic report or review has been published regarding the impact of this scheme. It is worth noting, however, that the role of the community champions is not far from what many mutual aid volunteers took it upon themselves to operate in the early days of the pandemic. As reported by Jones et al. (42), 57% of volunteers in mutual aid groups also supported their neighbours by providing information about the virus.

Moreover, Tiratelli and Kaye (25) distinguish between three types of local council approaches to community organisations and mutual aid groups: micromanaged, indifferent, and facilitative. In the micromanaged approach, councils seek to control the efforts of volunteers and community organisations, issuing orders in a prescriptive language of ‘should’ and ‘must’, an approach which has caused participants to view local government as an obstruction (25). In the indifferent approach, councils fail to support such groups and refuse to collaborate with them, an approach which potentially hobbles volunteering and damages public trust. For example, Locality (40) members have reported a lack of information sharing and joint planning, an approach which has led to duplication and confusion, as well as a lack of support in accessing funding. These two approaches are contrasted with the facilitative approach, in which local authorities find ways to support communities without smothering them (e.g., by providing practical help such as supplying mobile phones and card readers; proactively connecting volunteers with existing networks and other groups; or providing spaces and infrastructure to help groups organise) (25). In Bristol, the community hub Wellspring Settlement was able to develop a system with the local authority to have volunteers DBS-checked in 24 hours (44). As councils that have made concerted efforts in community engagement are the ones that have best facilitated their local mutual aid groups (25), councils should seek to give community organisations the freedom to operate whilst providing practical support and advice when needed.

### Consulting local communities

At this critical juncture it is crucial that government policy not be simply shaped by politicians, civil servants and scientists, but by local communities themselves (58). To our knowledge, the COVID-19 period has seen only two completed consultations of local communities thus far. These consisted of one by The West Midlands Combined Authority to guide its COVID-19 recovery (59), and one by the Scottish Government on the impact of COVID-19 on community organisations and their priorities for recovery (60). The panel for the West Midlands Combined authority agreed six priorities for the recovery: getting safely back to normality; ensuring clear guidance is provided as communities move out of ‘lockdown’; a strong healthcare system, including mental health; preparing children to go back to school in a supportive environment; creating new jobs and training with an emphasis on apprenticeships and entry-level jobs; promoting and supporting businesses, especially smaller and local businesses. Some of the priorities identified in the Scottish consultation involve supporting mental health; limiting the impact of future cuts and reduced services on communities; addressing employment issues; a low carbon recovery; tackling inequalities, and capitalising on the rise in community spirit (60). Bristol City Council are also currently conducting a multi-stage engagement process which will involve a survey, online forum, and citizen’s assembly regarding the city’s COVID-19 recovery, all of which will feed into its overall recovery plan (61).

Whilst consultation practices are important, it is also worth acknowledging their limitations. Tiratelli (62) argues that such forms of engagement are often merely a pro-forma procedure with non-intention of handing over power to the communities in any meaningful way. Such approaches need to be combined with a meaningful project of community mobilisation, which builds strong coalitions, leadership, and engenders local communities with the belief that they can enact real change (62). It is precisely this mass mobilisation which has proved so invaluable in a time of crisis, and if properly tended to may lead to even greater things.

## Conclusions

This review focused on the nature and dynamics of COVID-19 volunteering as well as on how community engagement was fostered during the COVID-19 pandemic in the UK.

In terms of what we have learned so far from COVID-19 volunteering, we conclude that COVID-19 models of volunteering were diverse, not only in terms of modes of organising (e.g., more or less horizontal, formal or informal) but also in terms of the activities that were developed (35, 36). Since the COVID-19 outbreak, communities across the country were able to mobilise and organise into multiple and diverse forms of community action and support. Some existing groups changed their focus and started to support their local neighbourhoods. People without previous experience of volunteering set up informal support groups from scratch in their local areas. Thousands of mutual aid groups were created, and many people were active helpers in providing information about COVID-19, shopping, packing and delivering food, fundraising and making donations, collecting prescriptions, dog walking, and offering emotional support through telephone helplines, among others (25, 35, 40). In terms of the profile of the volunteers, the studies reviewed suggest that the demographic makeup of COVID-19 volunteers partly reflected pre-existing trends and inequalities, by showing that women, working-age people, and middle-class people were more engaged in volunteering than other demographics (25, 37, 45, 49). In addition, higher levels of social support, cohesion and trust, and pre-existing social networks were important dimensions in explaining the emergence of COVID-19 volunteering and also the profile of volunteers (45, 50, 52). However, is still unclear whether these dimensions reflect the different profile of the volunteers or some consequences of engaging in volunteering during a crisis. Further research is needed to better understand the profile of volunteers during the outreach, as well as whether the demographic and geographic distribution of volunteering may simply reproduce and even reinforce the existing inequalities exacerbated by COVID-19. If so, any governmental response should address the underlying socioeconomic disadvantages which hinder effective voluntary action (50). Importantly, this review indicates that volunteering practices changed since the first UK ‘lockdown’ (23^rd^ March – June-July 2020), and that support groups adjusted their activities and actions over time (40). Overall, our review suggests that communities and groups began to reorient themselves beyond the temporally bound demands of the pandemic context, and towards more fundamental structural demands. We argue that this shows not only that groups are willing to continue to provide community support but that they are also able to change their focus and adapt to new needs and challenges. The ability to adjust and adapt the activities of the group is very promising, and future studies should look at COVID-19 volunteering patterns over time.

Furthermore, our review suggests that groups strategically developed activities to promote volunteer well-being and avoid overloading them with tasks (8, 53). Past research suggests that such strategies are likely to produce a positive effect, as a large amount of time devoted to volunteering activities is a predictor of burnout (63). A recent study has shown that people’s sense of community commitment is often a reason for sustained engagement and that cohesive community relationships are particularly relevant for continuous volunteering over time (30). Yet, and despite the extensive literature on the factors influencing participation in volunteering (64), the predictors of volunteering during the pandemic may be slightly different from others forms of volunteering (45). Hence, further research is needed to understand how COVID-19 community groups can be sustained over time, by examining the role of these structural, psychological and contextual variables.

Many factors were identified as fostering effective volunteering endeavours, including local knowledge, existing relationships and trust built up over the years by pre-existing organisations were crucial to enable effective large-scale responses (35, 50). Despite its successes and achievements, volunteer groups faced several challenges, with this review suggesting some important lessons on how to foster engagement with local communities during the COVID-19 period. In this regard, studies reviewed highlight the need for local councils to improve the way support is provided to volunteering groups and other community-based organisations (25). Such support is key for community engagement in pandemic conditions, and as a way to reduce contagion (17, 18, 20, 21). Further research should focus on how organisations, official and non-official bodies, can improve their collaboration and cooperation with volunteer-based groups, in order to foster community engagement and participation. Community champions schemes may have the potential to foster community engagement (6), but research is needed to understand the role of community champions and the role of official organisations in facilitating and fostering community-based volunteering during COVID-19. Additionally, providing more evidence on how communities can be effectively engaged in decision-making during and beyond this pandemic is also crucial. Considering the importance of public engagement and community support in pandemic conditions (17), such research is relevant for public health interventions far beyond the current COVID-19 pandemic.

## Data Availability

Not applicable

## Declarations

### Ethics approval and consent to participate

Ethical approval was not required.

### Consent for publication

Not applicable.

### Availability of data and materials

Not applicable.

### Authors’ contributions

GM and JD contributed to the conception and design of the review. GM performed data sources searches and analysed the material. GM and MFJ wrote the first draft of the manuscript. All authors contributed to the interpretation of data, reviewed, wrote and approved the final version of the manuscript.

## Acknowledgements

Not applicable.

## Footnotes

1 Someone who tested positive for COVID-19 must self-isolate for 10 days. If in contact with someone that has symptoms or has tested positive, the recommendation is to self-isolate for 14 days (10).

